# Declining Trend in the Initial SARS-CoV-2 Viral Load During the Pandemic: Preliminary Observations from Detroit, Michigan

**DOI:** 10.1101/2020.11.16.20231597

**Authors:** Said El Zein, Nivine El-Hor, Omar Chehab, Samer Alkassis, Tushar Mishra, Vichar Trivedi, Hossein Salimnia, Pranatharthi Chandrasekar

## Abstract

We report a downward trend in the initial SARS-CoV-2 viral load in nasopharyngeal swab samples of hospitalized patients with COVID-19 in Detroit, Michigan, coinciding with a decrease in the number of deaths during April-June 2020. A gradual decrease in the initial viral load reflected the downward progression of the pandemic.

## Introduction

As of November 13, 2020, more than 53 million cases of SARS-CoV-2 infection were reported worldwide. The majority of cases were detected in the United States with more than 242,000 deaths so far in the U.S. alone [1]. Few studies have evaluated the SARS-CoV-2 viral load (VL) dynamics in patients with different disease severities. Zheng S et al. demonstrated that patients with severe disease had late shedding peaks compared to patients with mild disease [2]. The authors also reported that the virus could be isolated up to 29 days after symptom onset with declining VLs over time [2]. More recently, Pujadas et al. in a correspondence, demonstrated that a high SARS-CoV-2 viral load at diagnosis is an independent predictor of mortality in a large hospital cohort[3]. Trends in initial or presenting viral load over time have thus far not been evaluated.

We describe a progressive downtrend in the level of initial SARS-CoV-2 VL detected in nasopharyngeal samples of hospitalized patients in Detroit, Michigan during April 4-June 5, 2020. We also report a significant association between the initial VL of hospitalized patients and their mortality.

## Methods

We conducted a retrospective study which included all patients who had a nasopharyngeal swab sample positive for SARS-CoV-2 infection by the Cepheid GenExpert instrument system – Rapid Real Time Polymerase Chain Reaction (RT-PCR) at the Detroit Medical Center during April 4, 2020-June 5, 2020. All sample containers, swabs, transport and storage, nucleic acid extraction and PCR protocols were similar for all included patients. This study included hospitalized patients as well as ambulatory patients. We included results of the initial PCR sample obtained on presentation and omitted any subsequent results obtained from the same patient. Medical records through July 20, 2020 were reviewed and all-cause in-hospital mortality was recorded. Weeks were divided into intervals of seven days starting from April 4, 2020. After May 9, 2020, we noticed a significant drop in the number of hospitalized SARS-CoV-2 cases, therefore, all cases between May 9, 2020 and June 5, 2020 were included in the 6+ week category.

Samples were considered positive for SARS-CoV-2 when the N gene (nucleocapsid) was detected. Although the RT-PCR test is qualitative and results are reported as positive and negative, the Exprt Xpress SARS-CoV-2 assay reports the cycle threshold (Ct) when the test is positive. The Ct value can be used to estimate the number of viral particles in a patient sample. Based on known information regarding the performance of real time PCR assays and the relationship between the Ct value and target concentration, every 3.3 decrease in Ct value indicates a log_10_ (10-fold) increase in target concentration in the sample. Using a quantified standard, we performed studies to determine the Ct value at different target concentrations. At 10,000 and 100,000 viral particles, the Ct values were at 43 and 39.4 respectively, showing a reduction of Ct values by 3.3 when the target concentration increased by one log_10_. Assuming a linear relationship between Ct value and target concentration, samples with Ct value of 26 and 36 should have viral concentrations of 10^9^ and 10^6^ respectively, a cutoff we used to delineate the intermediate VL category in our study. The lower limit of detection of our assay is around 250 genome copies/mL (95% confidence).

We used descriptive statistics such as medians and interquartile range to describe continuous variables and the Mann Whitney U test for data analysis. Based on our studies, we designated high, intermediate, and low VL samples to have a Ct value of ≤ 25, 26-36, and ≥ 37 respectively and used chi-square analysis to compare VL with survival status.

The Institutional Review Board reviewed the study protocol and ethical approval for the conduction of this study was granted under a waiver of informed consent (IRB No. RR19393). All relevant ethical guidelines have been followed.

The details of the IRB/oversight body that provided approval or exemption for the research described: IRB Administration Office at the Wayne State University School of Medicine

87 E. Canfield St, Detroit MI 48201

Lawrence Crane, MD – Professor of Medicine

Phone: +1-313-577-1628

## Results

A total of 708 nasopharyngeal swabs were analyzed from hospitalized patients (Figure 1A) and 282 swabs from ambulatory patients. Figures 1B and 1C represent the change in initial VL over time in ambulatory and hospitalized patients respectively. During the first week of the study (week of April 4, 2020), the initial VL in the PCR-positive respiratory samples was predominantly in the intermediate group for both hospitalized and ambulatory patients respectively (47.9%, n=123 and 45.5%, n=35). As the pandemic progressed, we noticed a progressive decline in the percent of positive samples in the high and intermediate VL categories with a concomitant rise in the percent of positive samples in the low VL category. By week five, 69% and 67% of the samples in both groups were in low VL category respectively. This trend coincided with a decrease in the percentage of deaths in hospitalized patients. Among hospitalized patients who died, the median Ct was 30.2 (n=179) compared to 37.1 for patients who remained alive (n=529) (p < 0.001) (Figure 1D). The outcome of illness of most ambulatory patients is not known, hence not included in the present study.

**Figure 1:**
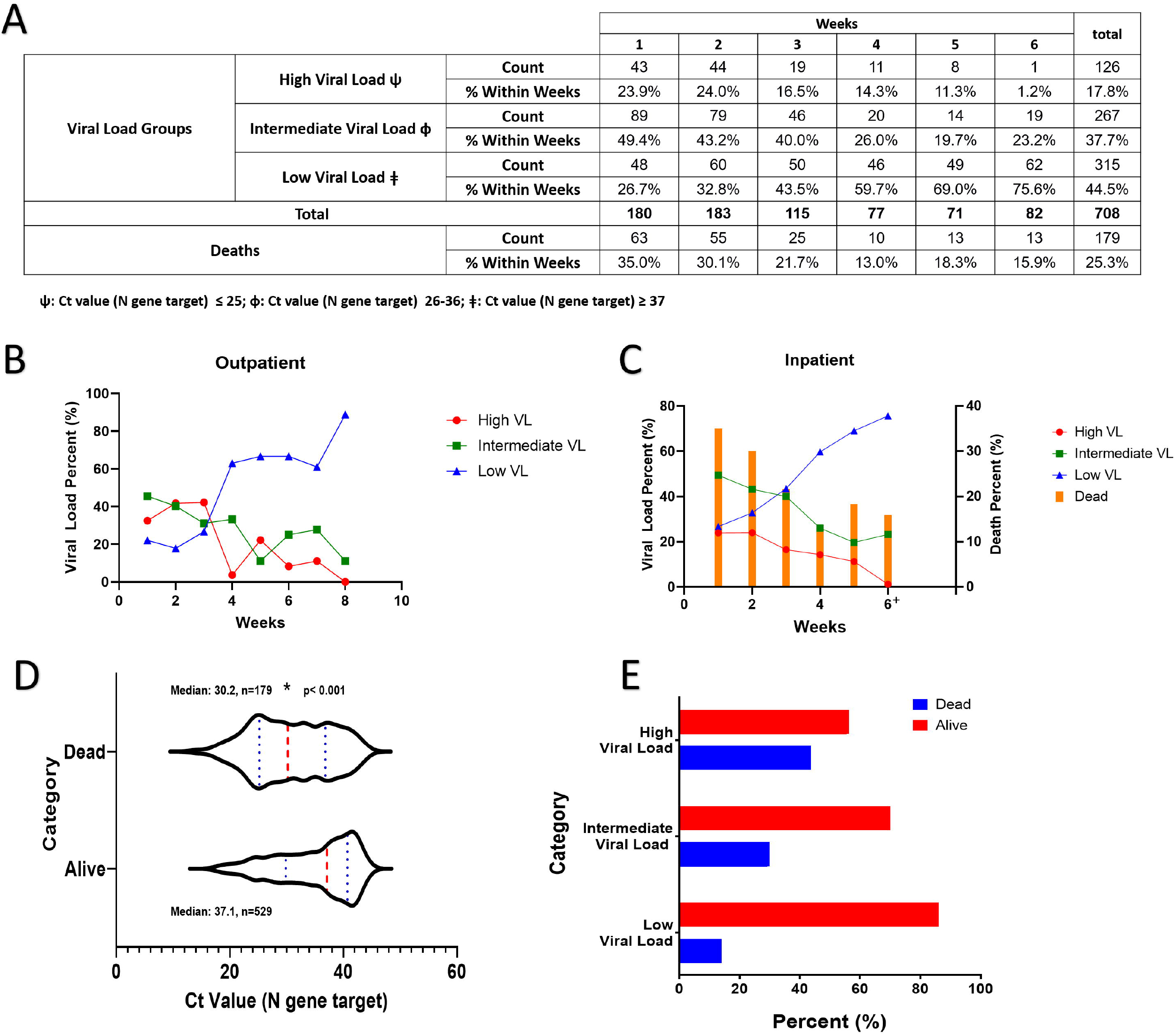
**A)** Table detailing the weekly number of SARS-CoV-2 PCR positive samples and respective within week percentages in each VL category in addition to the weekly number and within week percentage of reported deaths. **B, C)** Declining trend in the initial SARS-CoV-2 VL in ambulatory and hospitalized patients respectively. Weeks were divided into intervals of seven days starting from April 4, 2020. For hospitalized patients, weeks between May 9, 2020 and June 5, 2020 were included in the week 6+ category. **D)** Violin plot showing the distribution of the Ct values in hospitalized patients who were reported alive or dead during the course of the study. Red dots represent the median Ct value while blue dots represent the interquartile range. **E)** Bar graph representing the percentage of hospitalized patients reported dead or alive in each viral load category.

Figure 1E represents the relationship between VL and death among hospitalized patients. Almost one-half of the patients in the high VL category died compared to 30% and 14% in the intermediate and low VL categories respectively. There was a significant difference in the number of patients who died in the combined high and intermediate VL categories compared to patients in the low VL category (p < 0.001).

## Discussion

We observed that the initial SARS-CoV-2 load steadily declined among hospitalized and ambulatory patients during the April-June 2020 pandemic period, correlating with a decrease in the percent of deaths in hospitalized patients. Also, during this period, the aggressive nature of the pandemic gradually decreased. Exact reasons for the decrease in initial VL are unclear. Rapid implementation of social distancing measures and the widespread use of facemasks in Michigan may have contributed to less viral shedding and consequently, a lower viral inoculum on exposure. A change in the virus may also explain this observation, however, no data is available to support this conclusion. Tom MR et al, in an editorial, wrote that patients with a higher SARS-CoV-2 VL could be more infectious and suggested a testing framework to help determine which patients or exposed healthcare workers may have persistent transmissible disease [4]. Also, as the pandemic progressed, improved experience of the medical staff in managing COVID-19, patients seeking care earlier in their illness and more aggressive screening for COVID-19 in the emergency departments may have led to a lower initial VL among patients.

The correlation between VL, severity of illness and viral shedding remains a topic of debate. In patients with influenza infection, higher VLs were detected in hospitalized patients compared to ambulatory patients[5], however, this was generally not associated with worse outcomes [6]. In contrast, there is evidence that among patients infected with the Middle East respiratory syndrome coronavirus (MERS-CoV) and severe acute respiratory syndrome virus (SARS), a higher VL was an independent risk factor for death [7, 8]. Zheng S et al. reported that patients with more severe presentations had higher SARS-CoV-2 VLs [2], and recently, Magleby R et al. described an independent association between high VL, risk of intubation and in-hospital mortality [9]. Our data also suggest an association between initial VL and hospital mortality, though, other confounding variables may have influenced the findings. In contrast, Argyropulos K et al. describe an inverse relationship between SARS-CoV-2 VL, duration of symptoms and length of hospital stay. The authors found no association between VL and risk of intubation or mortality [10] and hypothesized that higher VLs are seen in milder disease.

This preliminary observational study has few limitations. First, the study does not include data about patient demographics, course of illness, duration of illness and comorbidities, which limit our ability to directly correlate higher viral loads with increased mortality. Currently, we are analyzing all variables and their impact. Second, we reported all-cause mortality and not deaths directly attributable to SARS-CoV-2 infection.

An upward trend in the initial Ct values (decreasing VL) of SARS-CoV-2 infected patients corresponded to a gradual attenuation of the severity of the pandemic. Positive behavioral modification such as social distancing and wider acceptance of facemask use may have contributed to a lower inoculum exposure, and hence a low initial viral load among the newly infected patients. Trends in the cycle threshold values over time may serve as a useful marker to assess the progress of the pandemic and may have an impact on public health measures, infection control and clinical management of infected patients.

## Data Availability

N/A

## Funding

No funding

## Conflict of Interests

The authors report no conflict of interests

## Notes

### Competing Interest Statement

The authors have declared no competing interest.

### Author Declarations

The Institutional Review Board reviewed the study protocol and ethical approval for the conduction of this study was granted (IRB No. RR19393). IRB Administration Office at the Wayne State University School of Medicine 87 E. Canfield St, Detroit MI 48201 Lawrence Crane, MD. Professor of Medicine Phone:+1-313-577-1628

## References

1. Centers for Disease Control and Prevention (CDC). Coronavirus Disease 2019 (COVID-19) 2020 [Available from: https://www.cdc.gov/coronavirus/2019-ncov/cases-updates/cases-in-us.html.

2. Zheng S, Fan J, Yu F, Feng B, Lou B, Zou Q, et al. Viral load dynamics and disease severity in patients infected with SARS-CoV-2 in Zhejiang province, China, January-March 2020: retrospective cohort study. BMJ. 2020;369:m1443–m.

3. Pujadas E, Chaudhry F, McBride R, Richter F, Zhao S, Wajnberg A, et al. SARS-CoV-2 viral load predicts COVID-19 mortality. Lancet Respir Med. 2020;[Epub ahead of print]. doi: https://doi.org/10.1016/S2213-2600(20)30354-4.

4. Tom MR, Mina MJ. To Interpret the SARS-CoV-2 Test, Consider the Cycle Threshold Value. Clin Infect Dis. 2020;[Epub ahead of print].

5. Granados A, Peci A, McGeer A, Gubbay JB. Influenza and rhinovirus viral load and disease severity in upper respiratory tract infections. J Clin Virol. 2017;86:14–9.

6. Lalueza A, Folgueira D, Muñoz-Gallego I, Trujillo H, Laureiro J, Hernández-Jiménez P, et al. Influence of viral load in the outcome of hospitalized patients with influenza virus infection. Eur J Clin Microbiol Infect Dis. 2019;38:667–73.

7. Feikin DR, Alraddadi B, Qutub M, Shabouni O, Curns A, Oboho IK, et al. Association of Higher MERS-CoV Virus Load with Severe Disease and Death, Saudi Arabia, 2014. Emerging infectious diseases. 2015;21:2029–35.

8. Chu C-M, Poon LLM, Cheng VCC, Chan K-S, Hung IFN, Wong MML, et al. Initial viral load and the outcomes of SARS. CMAJ. 2004;171:1349–52.

9. Magleby R, Westblade LF, Trzebucki A, Simon MS, Rajan M, Park J, et al. Impact of SARS-CoV-2 Viral Load on Risk of Intubation and Mortality Among Hospitalized Patients with Coronavirus Disease 2019. Clin Infect Dis. 2020;[Epub ahead of print].

10. Argyropoulos KV, Serrano A, Hu J, Black M, Feng X, Shen G, et al. Association of Initial Viral Load in SARS-CoV-2 Patients With Outcome and Symptoms. Am J Pathol. 2020:S0002-9440(20)30328-X.

